# Randomized Trial Protocol: Epic Generative AI Chart Summarization Tool to Reduce Ambulatory Provider Cognitive Task Load

**DOI:** 10.64898/2026.02.20.26346503

**Authors:** Aaron T. Chin, Nina Zhu, Thomas Kingsley, Pallavi Mynampati, Yan Phipps, Artem Romanov, Sitaram Vangala, Maxwell Weng, Lauren E. Wisk, Hawkin Woo, Ryo Isesu, John N. Mafi, Paul J. Lukac

**Affiliations:** UCLA Health Information Technology, UCLA Health, University of California, Los Angeles, Los Angeles, CA, United States; Department of Pediatrics, David Geffen School of Medicine, University of California, Los Angeles, Los Angeles, CA, United States; Division of General Internal Medicine and Health Services Research, David Geffen School of Medicine, University of California, Los Angeles, Los Angeles, CA, United States; David Geffen School of Medicine, University of California, Los Angeles, Los Angeles, CA, United States; RAND Corporation, Santa Monica, CA, United States; Department of Health Policy & Management, Fielding School of Public Health at UCLA; Department of Health Promotion and Human Behavior, Graduate School of Medicine, Kyoto University, Kyoto, Japan

**Keywords:** generative AI, EHR, chart review, clinician burden, task load, randomized trial

## Abstract

**Background:** EHR documentation and chart review contribute to clinician workload and burnout. To alleviate pre-charting burden, Epic has released a new generative AI chart summarizer tool, which has become widely adopted; however, its impact has not been examined in randomized trials.

**Objective:** To evaluate whether access to an Epic generative AI chart summarization tool reduces cognitive task load among ambulatory providers compared with usual care.

**Methods:** Two-arm, parallel-group randomized controlled trial among ambulatory clinicians across multiple specialties. Clinicians will be randomized 1:1 to tool access versus usual care for 90 days. The primary outcome is change in a 4-item physician task load (PTL) adapted for the pre-charting task. Exploratory outcomes include EHR-derived time metrics (Caboodle and Signal), professional fulfillment/burnout (PFI), usability (SUS), clinician satisfaction, aggregated patient experience item from CG-CAHPS, and reported safety related metrics.

**Ethics and Dissemination:** Analyses will use clinician-level survey responses and aggregated EHR metrics; no patient-level protected health information will be included in the analytic dataset. Results will be disseminated via preprint and peer-reviewed publication.

**Article summary – Strengths and limitations of this study:**

- This study is a 3-month pragmatic randomized controlled trial evaluating a native EHR-embedded generative AI tool that summarizes prior clinical notes for ambulatory encounters.
- The primary outcome uses a validated cognitive task load instrument adapted specifically for pre-charting activities.
- Exploratory outcomes include objective EHR-derived time metrics, validated psychometric measures of burnout and professional fulfillment, and clinician-reported survey measures assessing perceived usefulness of the tool.
- The trial is single-centered, which may limit generalizabilty, and the intervention is optional-use and unblinded, which may attenuate observed effects and introduce performance bias.

## Introduction

Clinical documentation in the electronic health record (EHR) is essential for sharing information among care teams, accurately tracking changes in a patient’s health, and supporting appropriate medical billing. However, significant documentation burden contributes heavily to physician burnout and is associated with increased medical errors, worsening workforce shortages and excess healthcare costs.(1-3) Excessive time spent interacting with the EHR, often extending into after-hours “pajama time”, is a major driver of this burnout.(4) In the ambulatory care settings, the common practice of “pre-charting” compels providers to review multiple aspects of a patient’s chart, such as recent notes and lab results, to contextualize the patient’s history with the reason for their visit.(5) This time-consuming process can impact quality, effectiveness, and efficacy of patient care.

Leveraging OpenAI’s GPT, Epic Systems, Inc [Verona, WI], an EHR company, has developed multiple summarization-based tools to ease administrative burden related to documentation and chart search and boost end user productivity. Across their suite of summarization tools, Epic generates over 16 million summaries monthly.(6) One such tool is a chart summary application that generates summaries of historical patient notes to assist ambulatory clinicians during pre-charting. One small pre/post quasi-experimental study on this functionality showed potential time savings with generally positive reception from physicians.(7) Despite this promise, deploying generative AI in healthcare introduces unique challenges. There is an inherent risk of hallucinations, inaccuracies, and omissions in AI-generated text, which could inadvertently increase physician documentation time and lead to clinical errors, among other things.(8-12) Additional challenges include technical limitations such as latency and scalability, as well as suboptimal human–AI interface design that may impede efficient integration into clinical workflows. As a result, the net value of these tools cannot be assumed and must be evaluated in real-world settings, particularly given the non-trivial financial, computational, and environmental costs associated with their deployment. These challenges underscore the importance of rigorous evaluations of generative AI (genAI) prior to and throughout their integration into clinical care.

Although Epic has released a rapidly expanding suite of genAI features, there remains lack of rigorous evaluation, including randomized controlled trials, to assess their usability, utility, or safety in real clinical workflows.(7, 13, 14) As a result, despite widespread interest, the broader healthcare community has little empirical evidence on how these tools perform in practice impeding informed purchasing decisions. To inform the design of this study, we drew on the methodology of a prior randomized controlled trial evaluating ambient scribes, which served as a valuable precedent.(9) By conducting a randomized controlled trial, this study will assist UCLA Health in making a data driven business decision, and will also help fill that knowledge gap by generating high-quality data on user experience and impact.

### Study Aims and Hypotheses

The primary aim of this study is to evaluate whether a native Epic genAI chart summarization tool reduces physician task load (PTL) compared to a control group. Exploratory endpoints include assessing the AI chart summarization tool on clinician metrics such as self-reported burnout, physician satisfaction, and productivity, as well as time efficiency with pre-charting tasks as measured via a Caboodle (native Epic dimensional database) metric.Additionally, we will explore the impact of this technology on the patient experience, specifically patients’ perceptions of how informed the clinician was about the patient’s medical history. Finally, we will evaluate whether prespecified baseline clinician and practice characteristics modify the effect of the tool.

## Methods

### Trial design

Two-arm, parallel-group randomized controlled trial with 1:1 allocation to intervention versus usual care control over a 90-day period.

### Setting

The study will be conducted in ambulatory outpatient clinics at the University of California, Los Angeles (UCLA) Health, an integrated academic health system using a single enterprise electronic health record (Epic Systems, Verona, WI). Multiple ambulatory specialties will be included to reflect routine clinical workflows.

### Participants

Eligible participants are ambulatory physicians and advanced practice practitioners (APP) across multiple specialties who hold at least one half-day of clinic per week. Participants will be recruited via email and/or survey invitation. Physicians in training are excluded.

### Inclusion criteria

- Ambulatory physicians and APP’s with at least one half-day clinic session per week
- Completes baseline survey

### Exclusion criteria

- Physicians in training (residents and fellows) and psychologists

### Randomization and allocation

A total of 284 ambulatory providers will be randomized 1:1 to the intervention or usual care control group. Randomization will be stratified by whether the participant has an active AI scribe license, and covariate-constrained randomization will be performed within strata to improve balance on baseline PTL (NASA-TLX–adapted score) and a modified baseline chart review time (Caboodle-derived). Due to the nature of the intervention, participants cannot be blinded to group assignment.

### Intervention

The trial intervention will span from 2/23/2026 - 5/24/2026, encompassing a 90-day study period. Participants in the intervention group will continue their usual clinical practice with access to Epic’s outpatient chart summarization tool. Use of the tool is optional and intended solely to provide a summary for providers and does not provide clinical decision support. It automatically generates a general or focused summary of the most recent clinical notes for any patient on a provider’s schedule. The number of notes summarized is limited by the character constraints of the EHR – 24,000 English characters or 30 notes. The tool includes a “focus on” feature allowing a short user prompt to center the summary on a specific topic. Users may also manually select notes for summarization. For scheduled patients, summaries are batch generated approximately 36 hours before the appointment; for same-day appointments, users may need to trigger summary generation. Summaries are created exclusively from clinical notes and do not draw from any other parts of the patient chart, such as labs and imaging, which will still require manual review by the physicians. The included note types will be progress notes, consults, procedures, H&P, discharge summary and ED provider notes. All participants will receive training before using the tool that encompasses tool functionality and general limitations/risks of genAI output.

### Comparator (usual care)

Clinicians in the control arm will continue usual pre-visit chart review workflows without access to the chart summarization tool during the 90-day period.

### Outcomes

#### Primary Outcome

Four-item physician task load (PTL), adapted from the NASA Task Load Index (TLX), is a validated tool for assessing EHR-related tasks using four sub-scales (mental demand, temporal demand, physical demand, and effort).(15) This outcome is adapted to capture the task of pre-charting, defined as the practice of reviewing patient information in the EHR prior to a visit. Each sub-scale is rated from 0 (low) to 100 (high) and is aggregated to a 0-400 point scale. No patient level information will be collected for this outcome measure.

#### Prespecified exploratory outcomes

| Outcome Measure | Measure Description |
| --- | --- |
| Modified Total Chart Time Per Encounter | Using Caboodle, Epic’s enterprise data warehouse, we will use a customized metric to measure clinician time spent reviewing the patient chart. Based on internal validation of where clinicians could access the chart summarization tool, the metric includes Caboodle Tier 1 activities corresponding to Clinical Review (all activities), Documentation limited to pre-charting activity only, and Other limited to Navigator-related activity, as well as the activity dedicated to the chart summarization tool. |
|  | All note-writing, order entry, and encounter-signing activities are excluded. Time is captured for office and telemedicine visits and includes pre-encounter and in-encounter activity, with no time captured after checkout. Change will be assessed relative to a 6-month retrospective baseline, with analyses focused on months 2 and 3. |
| Professional Fulfillment Index | The Professional Fulfillment Index (PFI) is a validated 16-item instrument that uses a 5-point Likert scale (0-4) to measure professional fulfillment, work exhaustion, and interpersonal disengagement.(16) Burnout is reported based on combined results of the work exhaustion and interpersonal disengagement subscales. |
| Self-Reported Pre-Charting Effectiveness | Providers will answer questions regarding self-reported effectiveness and efficiency in pre-charting. The questions use a 5-point Likert scale to measure these metrics, with a higher score indicating greater self-reported efficiency and effectiveness. |
| Provider Satisfaction Scores | Self-reported satisfaction survey that includes physician reported effect of chart summarization tool on pre-charting efficiency and effectiveness, and other potential unintended consequences. No patient level information will be collected for this outcome. |
| System Usability Scale | The system usability scale (SUS) is a ten-item questionnaire that uses a 5-point Likert scale to measure different aspects of system usability. The total scale ranges from 0-100. A higher SUS score indicates higher usability. |
| Reported Safety | Clinician-reported safety of AI-generated chart summaries, including perceived frequency of clinically significant errors and occurrence of major safety events during the intervention period. No patient level information will be collected for this outcome. |
| Tool-specific EHR feedback | Aggregated real-time feedback submitted by clinicians through the EHR-native summarization interface during the intervention period, including ratings of accuracy, completeness, and hallucinations. No patient-level information will be collected for this outcome. |
| Consumer Assessment of Healthcare Providers and Systems Clinician & Group Survey (CG-CAHPS) Metric | The Consumer Assessment of Healthcare Providers and Systems Clinician & Group Survey (CG-CAHPS) is a standardized patient feedback survey measuring experiences with providers.(17) We will examine changes in the CG-CAHPS mean scores compared to 6 months prior to the intervention for the question “In the last 6 months, how often did this doctor seem to know the important information about your medical history”. |
| Change in EHR Signal (Activity) Data - Time outside scheduled hours | We will examine change from a retrospective baseline 6 months prior to enrollment in Signal metrics including time outside scheduled hours per scheduled day. Using this data will determine how a provider’s time is utilized in the EHR. |
| Clinician RVUs per week | Average relative value units (RVUs) per week during intervention months 2 and 3, adjusted for baseline (6-month pre-intervention period). |

#### Effect Modification and Subgroup Analyses

The following subgroup analyses are prespecified and exploratory in nature, with a narrower focus on baseline clinician and practice characteristics.

- Baseline chart review time (Caboodle-derived; continuous).
- Clinician clinic volume or clinical FTE, defined using a prespecified baseline period; modeled continuously when feasible.
- Panel complexity (clinician-level RAF score; continuous).
- AI literacy and prior exposure to ambient AI scribes, including active AI scribe license status if available at baseline
- We will also conduct a prespecified exploratory causal machine learning analysis to evaluate heterogeneous treatment effects using baseline clinician, practice, and patient-panel characteristics available before treatment assignment.

#### Data Collection and Sources

The study dataset will include both pre-implementation and post-implementation data. Pre-implementation data will be collected via a Qualtrics survey administered prior to tool deployment to confirm physician eligibility, document opt-in, and capture baseline characteristics. Post-implementation data will be collected after three months of tool use and will include physician follow-up survey responses, EHR-derived usage metrics from Epic Caboodle and Signal, and aggregated patient encounter data. We will also extract aggregated, tool-specific feedback submitted within the chart summarization interface during the intervention period and calculate category-specific report rates relative to total summary generation volume. No patient-level protected health information will be collected; all EHR data will be de-identified, aggregated, and analyzed exclusively at the physician level.

#### Statistical Analysis

Linear mixed models will be used to estimate intervention effects, with provider random effects accounting for repeated measurements (baseline and follow-up for survey-derived outcomes; baseline, month 1, and months 2+3 for modified total chart time). Models will include fixed effects for study arm, response period, and the interaction of these terms, and will adjust for the following provider characteristics: provider sex, age, specialty (primary care, medical subspecialty, and surgical subspecialty), and number of half days in clinic. Intervention effects will be evaluated using linear contrasts between study arms in the follow-up period for survey outcomes, and in months 2+3 for modified total chart time. Exploratory analyses evaluating effect heterogeneity will be performed by evaluating interaction terms with the prespecified modifiers listed above and estimating effects in subgroups using linear contrasts. We will also perform an exploratory analysis contrasting the effect on modified total chart time in month 1 with that of months 2+3 using linear contrasts.

We will also conduct prespecified exploratory causal machine learning analyses to evaluate heterogeneity in the effects on ambulatory provider cognitive task load and exploratory outcomes, using causal forest models.(18) Causal forests estimate conditional average treatment effects (CATEs), defined as the expected treatment effect conditional on sample characteristics. Unlike traditional subgroup analyses, these algorithms can capture heterogeneity arising from high-dimensional, nonlinear interactions among multiple effect modifiers. Potential effect modifiers will include baseline clinician, practice, and patient-panel characteristics available before treatment assignment. To limit overfitting, we will use “honest” sample splitting, in which one subsample is used to grow the forest and a separate subsample is used to estimate CATEs.(19) We will apply 10-fold cross-fitting, and select tuning parameters via 10-fold cross-validation.(20) If potential effect modifiers are highly correlated and the causal forest models exhibit instability, we will reduce the set of potential effect modifiers included in the model to obtain more stable estimates. Samples will then be grouped into quantiles (e.g., quartiles) of predicted CATEs to summarize and compare sample characteristics across strata of heterogeneous treatment effect. As a sensitivity analysis, we will re-estimate heterogeneous treatment effects using Bayesian causal forests to assess robustness to modeling choices.(21)

The proposed intention-to-treat analysis of survey outcomes is robust to dropout under the missing at random (MAR) assumption. We will perform sensitivity analyses comparing the characteristics of responders and non-responders to assess the plausibility of this assumption, and will perform robustness checks using other approaches for addressing missing data (e.g., multiple imputation). Evaluation of the primary outcome will use an 0.05 significance level; all other analyses will report effect estimates and 95% confidence intervals without hypothesis testing. Analyses will be performed using R v. 4.5.1 (https://www.r-project.org/). The primary endpoint will remain the only confirmatory hypothesis test.

#### Sample Size and Power

The sample size of 284 participants provides 80% power to detect intervention effects as small as 0.33 standard deviations (small-to-medium effect size). This assumes a two-sample t-test on the pre-post change (a conservative approximation of the planned linear mixed model analysis), and a two-sided 0.05 significance level.

#### Data monitoring and missing data

Given the minimal-risk, clinician-facing nature of the study and absence of patient-level outcomes, no data monitoring committee is planned. EHR-derived metrics from Caboodle and Signal are expected to be complete for enrolled clinicians so missing data for these outcomes is expected to be negligible. For survey-based measures, we expect high follow-up survey completion but we will evaluate the extent and source of missing by comparing all baseline characteristics for clinicians with complete vs incomplete data at follow-up – allowing us to assess the plausibility of a MAR assumption. Subsequent missing data strategy for follow-up survey data will be informed by this evaluation.

#### Patient involvement

Patients were not involved in the design, conduct, reporting, or dissemination plans of this research. The study focuses on clinician-facing workflow tools, and outcomes are measured at the clinician level using survey instruments and aggregated EHR-derived metrics.

#### Ethics approval and consent

IRB-26-0066, approved. All participants will provide informed consent prior to enrollment.

#### Privacy and data security

Data will be stored on secure institutional systems with access restricted to study personnel. The analytic dataset will be limited to clinician-level survey responses and aggregated clinician-level EHR metrics.

## Discussion

Our study aims to evaluate the impact of a an EHR developed generative AI chart summarization tool on cognitive task load in the ambulatory setting through a two-arm RCT over a 3-month period. Results from this study will inform UCLA Health business decision-making and also explore how a generative AI note summarization tool improves physician pre-charting time and reduces burnout.

There are several limitations to this study. The three-month time period is relatively short for the intervention to adequately capture long-term effects of a chart summarization tool. Physician workload also varies by season as well as ranging from full day to half-day clinics, which may also impact ability to familiarize themselves with the tool. Another limitation is that participation is voluntary, limiting generalizability to providers who may be less inclined to engage with new technologies. Furthermore, this study will be conducted within a single academic health system using a specific EHR platform, which may limit the applicability of the results to other healthcare settings or systems.

This study may be subject to several potential biases. First, participants will not be blinded to their assignment. The reliance on self-reported surveys for key secondary outcomes, such as physician burnout, introduces subjectivity that may not fully reflect the impact of a chart summarizing tools. Additionally for users that use the tool but interact with it minimally, usage may not be reflected as accurately, as usage is tracked based on clicking on items within the tool.

To our knowledge, this is the first RCT evaluating the impact of an EHR-developed generative AI tool. These findings will provide insights into its effect on efficiency, productivity and physician well-being in ambulatory care settings.

## Trial Registration

ClinicalTrials.gov registration submitted Feb 20, 2026, and pending review at the time of submission.

## Data availability statement

The datasets generated and analyzed during this study will not be publicly available due to institutional restrictions and the use of internal EHR-derived metrics. De-identified, aggregated data may be made available from the corresponding author upon reasonable request and subject to institutional approvals.

## Ethics Statement

This study was approved by the UCLA Institutional Review Board (IRB-26-0066). No patient-level data will be collected.

## Author contributions

All authors contributed to the conception and design of the study, reviewed the manuscript, and approved the final version.

